# Validating Field-Feasible Measures of Recent Khat Use: A Diagnostic Accuracy Study Comparing Amphetamine Immunoassay and Assisted Self-Report Against HPLC in an Ethiopian Male Cohort

**DOI:** 10.64898/2026.06.14.26355466

**Authors:** Hugh Atkinson, Zeleke Mekonnen, Sultan Suleman, Laura Oetzel, Matiwos Soboka, Marina Widmann, Stefan W. Toennes, Markos Tesfaye, Samuel A. Stewart, Veronika Müller-Bamouh, Thomas G. Schulze, Mark Asbridge, Manuel Mattheisen, Kristina Adorjan, Michael Odenwald

**Affiliations:** Department of Community Health and Epidemiology, Dalhousie University, Halifax, Canada; Department of Medical Laboratory Sciences, Faculty of Health Sciences, Jimma University, Jimma, Ethiopia; Jimma University Laboratory of Drug Quality (JuLaDQ) and School of Pharmacy, Jimma University, Jimma, Ethiopia; Department of Psychology, University of Konstanz, Konstanz, Germany; Department of Psychiatry and Psychotherapy, University Hospital of Psychiatry, University of Bern, Switzerland; Department of Psychology and Neuroscience, Dalhousie University, Halifax, Canada; vivo international e.V., Germany; Goethe University Frankfurt, University Hospital, Institute of Legal Medicine, Frankfurt Main D-60596, Germany; Department of Psychiatry and Behavioral Sciences, State University of New York Downstate Health Sciences University, Brooklyn, NY, USA; Institute for Genomics in Health, State University of New York, Downstate Health Sciences University, Brooklyn, NY, USA; College of Pharmacy, Dalhousie University, Halifax, Canada; Institute of Psychiatric Phenomics and Genomics (IPPG), Medical Center of the University of Munich, Germany; Department of Psychiatry and Behavioral Sciences, SUNY Upstate Medical University, Syracuse, NY, USA; Department of Psychiatry and Behavioral Sciences, Johns Hopkins University School of Medicine, Baltimore, MD, USA; German Center for Mental Health (DZPG), partner site Munich-Augsburg, Munich, Germany; Department of Emergency Medicine, Dalhousie University, Halifax, Canada Faculty of Computer Science, Dalhousie University, Halifax, Canada; Africa-Europe CoRE in Non-Communicable Diseases & Multimorbidity, African Research Universities; Alliance (ARUA) & The Guild of European Research-intensive Universities, University of Bern, Switzerland

**Keywords:** khat, Catha edulis, diagnostic accuracy, STARD, self-report, immunoassay, HPLC, Ethiopia, substance use measurement

## Abstract

**Background:** Khat (*Catha edulis)* is a widely consumed natural amphetamine-analog used across East Africa and the Arabian Peninsula. Accurate field-feasible measurement of recent khat use is a prerequisite for large-scale epidemiological research; yet no validated alternatives to laboratory reference methods have been identified in the scientific literature. This nested validation study evaluated the diagnostic accuracy of two point-of-care measures, a commercial amphetamine immunoassay and a Timeline Followback (TLFB) Assisted Self-Report (ASR), against high-performance liquid chromatography (HPLC) quantification of urinary norephedrine (NE), while additionally assessing agreement between the two field measures.

**Methods:** A prospective, random sub-sample of 119 male participants aged 18-40 years from the Gilgel Gibe Field Research Center (GGFRC) longitudinal cohort, Ethiopia (validation timepoint T2, 2015), was used. Three index-reference comparisons were conducted: (1) amphetamine immunoassay (nal von minden, Drug-Screen AMP test, 300Öng/mL cutoff) vs.ÖHPLC; (2) binary ASR (past-week use) vs.ÖHPLC; and (3) binary ASR vs.Öimmunoassay. Sensitivity (positive percent agreement, PPA), specificity (negative percent agreement, NPA), positive predictive value (PPV), negative predictive value (NPV), overall accuracy (overall percent agreement, OPA), and Cohen’s kappa were calculated with 95% confidence intervals. Pre-specified secondary analyses applied three pharmacokinetically-informed recall windows (0-2, 3-5, and 6-7 days prior to interview) to ASR.

**Results:** Against HPLC (77 positive, 42 negative), the immunoassay showed perfect specificity (1.0 [0.916-1.0]) and PPV (1.0 [0.91-1.0]) but low sensitivity (0.52 [0.40-0.64]), NPV (0.53 [0.42-0.65]), overall accuracy (0.69 [0.60-0.77]), and weak kappa (0.43 [0.34-0.52]). Binary ASR showed high sensitivity (0.96 [0.89-0.99]), specificity of 0.60 [0.433-0.74], PPV (0.81 [0.72-0.89]), NPV (0.89 [0.72-0.98]), with overall accuracy 0.83 [0.75-0.89] and moderate kappa (0.60 [0.51,0.69]). Restricting ASR to use within 0-2 days improved specificity to 0.69 [0.52-0.84], PPV to 0.86 [0.77-0.93], overall accuracy to 0.87 [0.79-0.93], and kappa to 0.69 [0.61-0.78] (moderate), while sensitivity (0.96 [0.89-0.99]) and NPV (0.89 [0.72-0.98]) remained stable. Against the immunoassay, ASR achieved high PPA of (1.0 [0.91-1.0]), NPA of 0.35 [0.25-0.47], OPA of 0.57 [0.48-0.66], and minimal kappa (0.27 [0.19-0.35]).

**Conclusions:** Time-stratified ASR (0-2 days) is a valid, scalable alternative to biological testing for recent khat use in resource-limited settings. The immunoassay’s 300 ng/mL cutoff functions as a marker of heavy or recent high-dose khat use rather than any-use detection. Its perfect specificity and PPV make it valuable as a confirmatory test for substantial exposure, while its lower sensitivity reflects calibration to amphetamine rather than to khat-derived cathinone metabolite.

**Registration:** Not registered.

## Introduction

Khat (*Catha edulis)* is a flowering shrub widely cultivated and consumed across East Africa and the Arabian Peninsula^1^. Its leaves are chewed for their stimulant properties, principally attributed to the alkaloid cathinone and its metabolites^2^. In Ethiopia, national surveys estimate khat use prevalence exceeding 45% among adults in high-consumption regions^3^. Despite this, the epidemiology of khat use and its associated harms remains poorly characterised, in part because validated, field-deployable measures of recent use are lacking^4^. This measurement gap limits the field’s capacity to characterise both prevalence trends and exposure-outcomes associations in khat-using populations.

Several approaches exist for measuring recent khat use. Laboratory-based quantification of cathinone and its metabolites in biological samples represents the analytical reference standard. While whole-blood gas chromatography-mass spectrometry (GC-MS) of parent cathinone offers the highest analytical precision, it requires invasive sampling, cold-chain transport, and specialised laboratory infrastructure that are rarely feasible in field-based research in low-and-middle-income countries (LMICs). Urinary high-performance liquid chromatography (HPLC) quantification of norephedrine (NE), the primary stable urinary metabolite of cathinone, provides a non-invasive alternative with strong analytical validity and was used as the reference standard in this study^5^. However, even methods like urinary HPLC require specialised infrastructure and trained personnel, making it expensive and logistically impractical for large community-based studies in LMICs. Commercial immunoassay strip tests (e.g. to detect amphetamine-class substances) offer a low-cost, rapid biological alternative, but have not been validated specifically for detecting khat-derived alkaloids at standard cutoffs.

Although both amphetamine-class immunoassay and Timeline Followback (TLFB) Assisted Self-Report (ASR) have established performance profiles for amphetamine-type stimulants, opioids, and cannabis in the broader substance use literature, their accuracy for khat-derived alkaloids specifically has not been established.

TLFB ASR is logistically feasible and leverages existing community surveillance networks but is subject to recall bias and social desirability effects. A further methodological complication for ASR is temporal misalignment with biological detection. Urinary amphetamine has a maximum detection window of about 50-hours post-ingestion, while urinary NE remains detectable for approximately 3 days following use^5–7^. Inter-individual variation in the CYP450 activity, which khat use has been shown to inhibit, may further affect cathinone clearance rates and thus the effective detection window in practice^8^. Self-reported use beyond this window therefore cannot be confirmed biologically, producing apparent ASR-Immunoassay discordance that is pharmacokinetically driven, rather than behaviourally. Stratifying self-report into pharmacokinetically-informed time windows is potentially an important analytical strategy for improving the interpretability of ASR accuracy.

The intended use of the measures evaluated here is epidemiological surveillance: large-scale, repeated cross-sectional or longitudinal assessment of khat use in community populations where laboratory testing is unavailable. In this context, the immunoassay is evaluated as a potential replacement for sophisticated lab-based methods as a biological field standard, and ASR is evaluated both as a replacement for the two biological approaches when even point-of-care testing is not feasible.

Study objectives and hypotheses: This study has three pre-specified objectives:

1. To determine the diagnostic accuracy (sensitivity, specificity, positive predictive value (PPV), negative predictive value (NPV), overall accuracy, and kappa) of the nal von minden, amphetamine immunoassay (300Öng/mL cutoff; AMP300) against HPLC as the reference standard. We hypothesised that the immunoassay would show high specificity for amphetamine class compounds but reduced sensitivity relative to HPLC, given its detection threshold.
2. To determine the diagnostic accuracy of binary TLFB ASR (past-week khat use) against HPLC as the reference standard, including pre-specified secondary analyses using pharmacokinetically-informed time windows (0-2, 3-5, and 6-7 days prior to interview). We hypothesised that ASR would show high sensitivity but reduced specificity relative to HPLC, with improvement in specificity when restricted to shorter recall windows aligned with the NE detection window.
3. To assess the level of agreement between binary ASR and the immunoassay as co-deployed field measures, to determine whether ASR can function as a stand-alone substitute for immunoassay testing in large-scale community research.

## Methods

### Study Design

This is a secondary analysis of data from a prospective longitudinal cohort study conducted at the Gilgel Gibe Field Research Center (GGFRC), Ethiopia. The original study, examined the prevalence and associations of substance use and common mental disorders among young men in the Gilgel Gibe Field Research Center, Ethiopia, led by investigators at Ludwig Maximilian University of Munich (LMU), the University of Konstanz, and Jimma University, Ethiopia^9^. The diagnostic accuracy analyses reported here, comparing two index tests (an amphetamine immunoassay and TLFB ASR) against urinary HPLC as the reference standard, use data from a pre-planned validation timepoint (T2) nested within that parent study, with index test and reference standard data collected concurrently and prospectively. The three primary comparisons and the time-stratified secondary analyses were all pre-specified prior to conducting any statistical analyses. Ethical approval was granted by the Jimma University Institutional Review Board (Ref: RPGC/546/2014 and Ref: IHRPGD/2069/2017) and the LMU Institutional Review Board. Written informed consent was obtained from all participants in their preferred language (Afaan Oromo or Amharic). Data protection measures included coded participant identifiers with linking logs stored separately, encrypted digital storage, and access restricted to named research personnel. This manuscript was drafted in adherence to the Standards for Reporting Diagnostic Accuracy (STARD 2015) reporting guidelines for diagnostic accuracy studies^10^.

### Participants

#### Parent cohort and setting

The GGFRC is a Health and Demographic Surveillance System (HDSS) located in the Jimma Zone, south-western Ethiopia, covering 11 kebeles (3 urban, 8 rural) across four woredas within approximately 10 km of the Gilgel Gibe Hydroelectric dam, situated around 260 km south-west of Addis Ababa^11,12^. More than 60,000 residents, predominantly of Oromo ethnicity, live within the catchment area. The GGFRC has operated as a comprehensive surveillance site since 2005, monitoring vital events through biannual population updates and periodic censuses, and is a member of the INDEPTH Network of HDSSs in LMICs^11,13^. Sixteen local enumerators employed by Jimma University, fluent in both Afaan Oromo and Amharic, conducted field data collection within the kebeles where they reside. For the parent study, they received specialised training from psychiatrists, psychologists, and psychiatric nurses following a task-shifting model. Baseline data collection (T1) took place during February-March 2015 (the dry season), a period of restricted khat supply^14^. The final follow-up (T3) was conducted December 2015-January 2016, at the tail-end of the rainy season when khat was more readily available, approximately nine months after T1 (mean (SD) = 285 (10.3) days)^14^.

#### Eligibility criteria and validation sub-sample

Eligibility criteria for the parent cohort were male sex, age 18-40 years, residence within the GGFRC catchment area for at least six months, ability to provide informed consent, and no severe cognitive impairment preventing interview completion (for example, active epilepsy). Of 1,100 initially screened, 259 declined consent, yielding a final T1 sample of 841 participants.

Participants were identified through the GGFRC biannual enumeration system: an additional question was added to the routine household census asking whether residents would be willing to participate in the study, and eligible respondents in the target age range were selected using stratified random sampling by kebele.

At T2 (conducted on average 3 (SD 1.9) days after T1 at a local health centre within walking distance of participants’ homes), 126 participants were randomly selected from the 841 eligible T1 respondents. T2 assessments were conducted by expert enumerators (psychiatrists, psychologists, and psychiatric nurses), unlike T1 and T3 which were conducted by trained lay enumerators at participants’ homes. Participants were excluded from the diagnostic accuracy analyses if they reported use of over-the-counter cold medications in the preceding week (nÖ=Ö4; such medications may contain pseudoephedrine or phenylpropanolamine, which are amphetamine analogues detectable by the immunoassay and may confound HPLC NE quantification) or had invalid or missing HPLC results (nÖ=Ö3). The final analytic sample comprised 119 participants.

**Figure 1.**
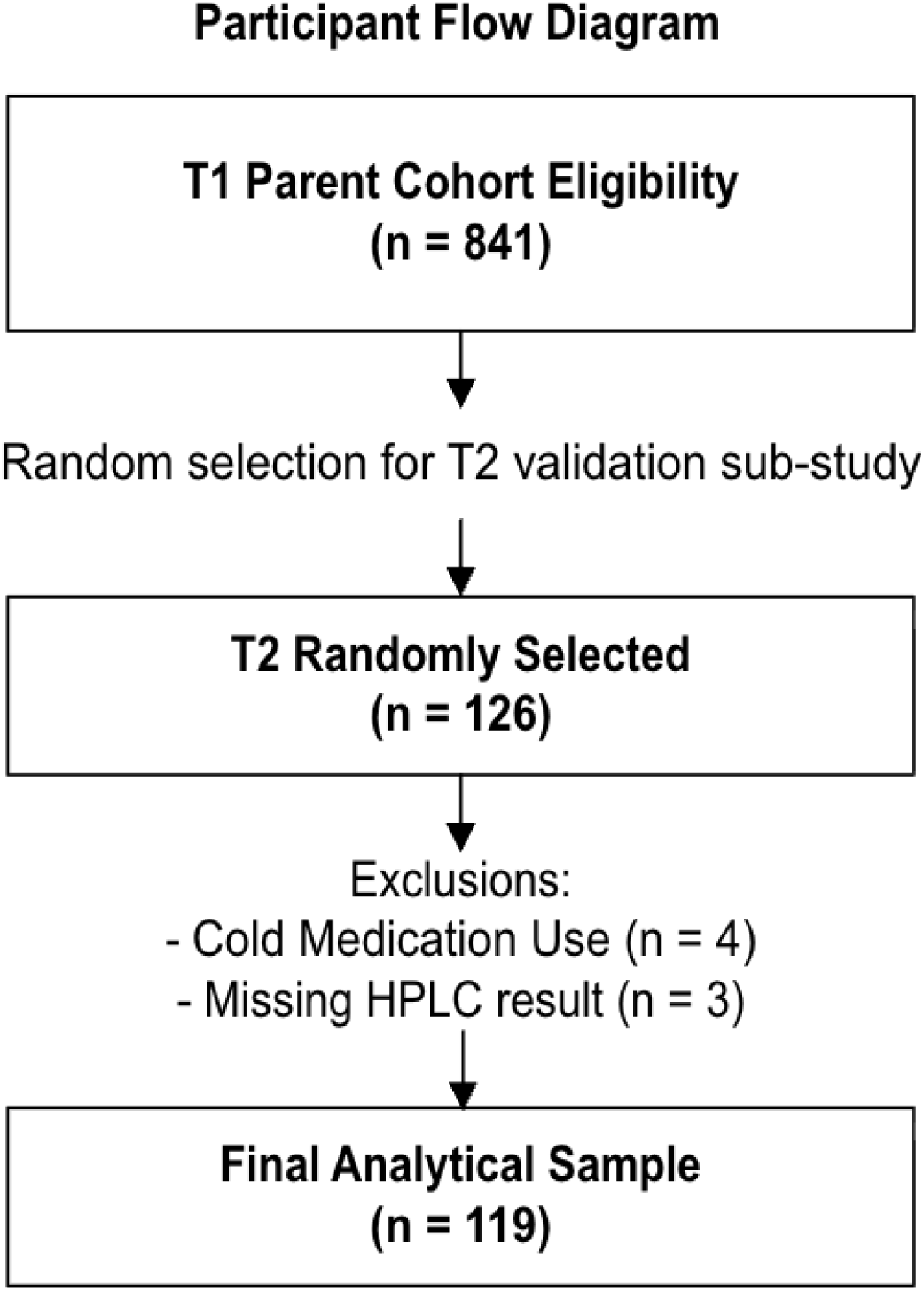
Participant flow.

### Measures

#### Reference standard: HPLC quantification of urinary NE

Urine samples were collected at T2 in labelled standard containers. Samples were refrigerated at local health centres and transported to Jimma University in cold boxes to maintain temperature control, where they were stored at 2-8°C until analysis at the Jimma University Laboratory of Drug Quality (JuLaDQ) and School of Pharmacy, Jimma University, Jimma, Ethiopia. HPLC was used to quantify urinary NE concentration. NE is both a minor constituent alkaloid of khat and the primary stable urinary metabolite of cathinone, the principal psychoactive alkaloid in khat, and was selected for analysis by the original collection team due to its metabolic stability (remaining detectable for more than one day after use) and its legal status in Ethiopia. NE is also analytically stable under field storage conditions: stability testing indicates that urinary NE remains stable at temperatures up to 20°C for at least 14 days^15^. Given the samples were refrigerated between 2-8°C and reached the laboratory within a mean (SD) of 4.9 (1.7) hours, degradation during transport is unlikely to have materially affected NE quantification. HPLC was selected as the reference standard over alternative methods such as gas chromatography, which requires more complex sample preparation, on the basis of its superior precision, reliability, and ability to provide exact NE concentration values.

Reference standard positivity threshold: Because NE is an exogenous compound that is not endogenously produced, its presence in urine indicates khat exposure. Urinary NE was quantified by HPLC as a continuous concentration; a positive result was defined as any concentration above zero. All 42 non-detects read exactly 0 µg/mL and the lowest quantified concentration was 1.8 µg/mL, with no intermediate values, so HPLC-positive status corresponds to unambiguous NE detection. A result was classified as positive if urinary NE concentration exceeded the analytical limit of detection of the HPLC method (the lowest concentration distinguishable from a blank). Details of this calculation are available from the corresponding author on request. For analytical purposes, any detectable NE above this threshold was treated as a positive indicator of recent khat use. This threshold was pre-specified. The rationale is that any detectable NE at or above this limit represents biological evidence of recent phenylpropanolamine isomer exposure, since NE has no endogenous source. While HPLC provides the most precise available measure of recent khat exposure, it is acknowledged that inter-individual variability in metabolite excretion may introduce some measurement error. Full details of the HPLC method development and validation are reported separately (Adorjan et al., in preparation).

#### Index test 1: Amphetamine immunoassay

The nal von minden, Drug-Screen AMP test (300 ng/mL cutoff, Moers, Germany), a urine-based amphetamine immunochromatographic strip test, was selected based on systematic pretesting and used to assess the presence of khat alkaloids in participants’ urine.

This lateral-flow competitive immunoassay is calibrated to detect amphetamine; cathinone and its metabolites are detected through structural cross-reactivity with the amphetamine antibody rather than direct binding. Immunoassay tests were administered at all three time points (T1, T2, T3) by trained pharmacological assistants or nurses at local health centres, using the same urine sample collected for HPLC at T2. Tests were performed on the day of collection following the manufacturer’s protocol^16^. A key practical advantage of the immunoassay is that it requires no electricity and can therefore be deployed at rural health centres without laboratory infrastructure, making it suitable for resource-limited field settings.

Index test positivity threshold: A positive result was defined as an amphetamine concentration exceeding 300Öng/mL, per the manufacturer’s standard cutoff. This cutoff was pre-specified and not modified for this study.

#### Index test 2: Assisted Self-Report (ASR)

Khat use was assessed using a modified version of the Timeline Followback (TLFB) approach, administered as a structured face-to-face interview at T1, T2, and T3^17^. The TLFB is a calendar-based retrospective recall instrument that elicits day-by-day substance use over a defined reference period using a calendar to assist recall^17^. At T2, expert enumerators (psychiatrists, psychologists, or psychiatric nurses) administered the TLFB, covering the day of interview (day 0) and the seven preceding days (days 1-7), an 8-day window. For each day, the instrument recorded: whether khat was used (yes/no), total hours of use per day, start and stop times, the type of khat, and the monetary value of khat consumed. Summary measures included total days chewed, total hours spent chewing in the preceding week, and time between urine collection and last khat consumption. Participants with no khat use in the prior 8 days were assigned 192 hours (the 8-day ceiling); these values are right censored, not measured.

Binary ASR variable (primary, pre-specified): Any self-reported khat use across the full 8-day TLFB window (i.e., including the day of interview itself) was coded as positive. This variable was derived from the TLFB day-by-day grid rather than from a single summary question (see Appendix A).

Time-stratified ASR variables (secondary, pre-specified): Three pharmacokinetically-informed recall windows were pre-specified in the analysis plan prior to any statistical analysis, based on the known urinary NE excretion profile following khat use: (a) most recent use 0-2 days prior to interview (very recent use, within the window of peak urinary NE concentration); (b) most recent use 3-5 days prior (alkaloid decline phase, where khat alkaloids begin to degrade but remain detectable); (c) most recent use 6-7 days prior (extended recall window, expected to be beyond the biological detection window)^7,18,19^. These windows allow investigation of how temporal alignment between self-report and biological detection affects diagnostic accuracy.

#### Test independence and blinding

To minimise bias, HPLC analysis was performed by laboratory technicians at the Jimma University Laboratory of Drug Quality who were blinded to both immunoassay results and ASR khat-use status at the time of analysis. Samples were transported and logged with coded identifiers only. Similarly, immunoassay test performers at local health centres did not have access to participants’ ASR khat-use data at the time of testing. ASR interviews at T2 were conducted by expert enumerators prior to immunoassay results being available. It is acknowledged that, due to the nature of the study, HPLC analysts could not be fully blinded to the fact that samples originated from a khat use study; however, this does not represent directional contamination of individual results. There was no cross-contamination of results between the immunoassay, HPLC, and ASR at the time of data collection.

### Statistical Analysis

#### Diagnostic accuracy estimation

For each index-reference comparison, a 2×2 contingency table was constructed yielding true positives (TP), true negatives (TN), false positives (FP), and false negatives (FN) against the binary HPLC outcomes as reference standard. The following metrics were calculated with 95% confidence intervals using exact binomial methods:

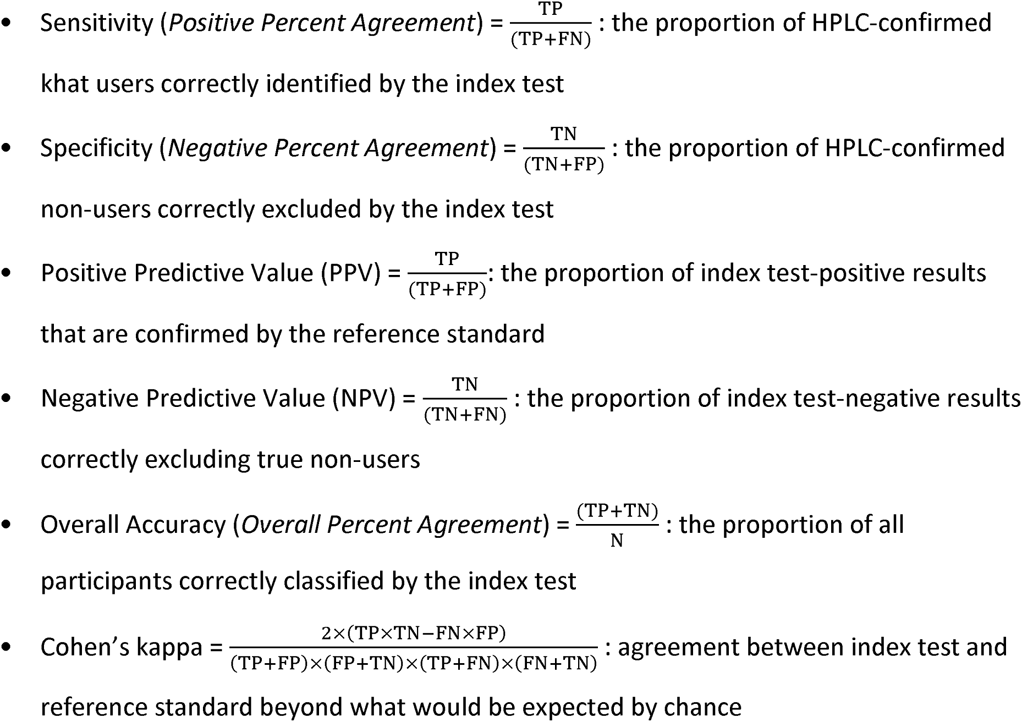

**Figure 2.**
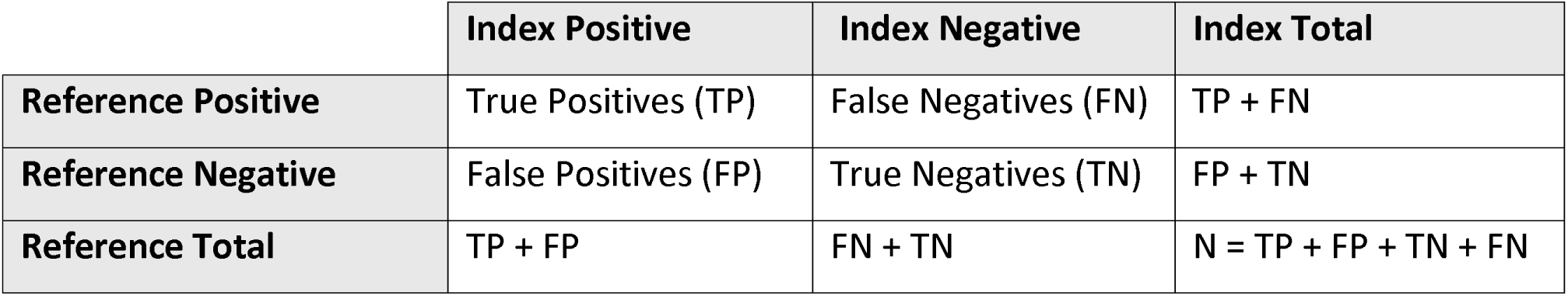
Sample 2×2 contingency table

Cohen’s kappa was calculated for each comparison to assess agreement beyond chance, with values interpreted using standard intervals^20^. Given the observed prevalence imbalance in the validation sample (∼76% khat users by ASR), kappa was treated as a primary metric as it is less sensitive to prevalence than overall accuracy. For comparisons lacking a validated reference standard, Positive Percent Agreement (PPA), Negative Percent Agreement (NPA) and Overall Percent Agreement (OPA) will be reported in place of sensitivity, specificity, and overall accuracy. Reporting guidelines for diagnostic validation studies mandate these terms, as specificity and sensitivity assume a validated reference standard^21^. Predictive values will not be reported for these comparisons. Discordant pairs were examined to characterise plausible sources of error. All analyses were conducted in R (version 4.5.1).

To further explore the validity of the immunoassay and the binary ASR, independent-samples t-tests (Welch’s) were conducted comparing quantitative NE concentrations between concordant and discordant classification groups. Specifically, for each index-reference comparison, mean NE concentration was compared between (a) HPLC-positive participants correctly identified by the index test (true positives) and (b) HPLC-positive participants missed by the index test (false negatives). These comparisons test whether misclassification is associated with lower analyte concentration, which would support a threshold-related, rather than random, explanation for false-negative results.

To further investigate the dose-dependent nature of immunoassay detection, total hours of khat use in the preceding 48 hours and across the full recall window were compared between immunoassay-concordant (ASR-Positive and immunoassay-positive) and immunoassay-discordant (ASR-positive but immunoassay-negative) participant among all 91 ASR-positive individuals. Mean hours were compared using Welch’s independent-samples t-tests, with effect sizes reported as Cohen’s d.

#### Indeterminate and missing results

No indeterminate immunoassay results occurred in this sample; all immunoassay tests produced a valid, interpretable result per the company’s criteria. Complete ASR and immunoassay data were available for all 119 participants in the final analytic sample; no missing data imputation was required for any variable.

#### Pre-specified and exploratory analyses

The three primary diagnostic accuracy comparisons (immunoassay vs. HPLC; binary ASR vs. HPLC; binary ASR vs. immunoassay) and the three pharmacokinetically-informed time-stratified secondary analyses (0-2, 3-5, and 6-7 days) were all specified in the analysis plan prior to conducting any statistical analyses of these data. Upon review, the 6-7 day stratified group contained only three participants (n=3), and it was therefore decided that the 3-5 and 6-7 day windows should be collapsed into a single 3-7 day group to protect participant confidentiality and ensure satisfactory cell counts for stable estimation of diagnostic metrics. No other exploratory post-hoc analyses are reported in this paper. The descriptive comparison of binary versus time-stratified ASR performance metrics (absolute change in sensitivity, specificity, predictive values, and kappa) is a pre-specified secondary analysis designed to quantify the effect of recall window alignment on diagnostic performance.

### Sample size

The validation sub-sample size (nÖ=Ö126, final analytic nÖ=Ö119 after exclusions) was determined based on resources available for HPLC laboratory analysis at T2 and the feasibility of calling back participants after baseline (T1). An a priori power calculation using exact binomial (Clopper-Pearson) intervals indicated that, assuming sensitivity of 0.80, the minimum threshold classified as ‘good’ diagnostic accuracy in substance use measurement, with 77 HPLC-positive participants, the expected 95% CI would be (0.72,0.88), and assuming specificity of 0.80 with 28 HPLC-negative participants, the expected 95% CI would be (0.70, 0.89)^22^. Assuming a specificity of 0.80 across the 42 HPLC-negative participants the expected 95% CI would be approximately (0.66,0.91). These bounds confirm that the sample is sufficient for estimating diagnostic accuracy and inspecting discordant pairs, though not for producing the narrow confidence interval width required for policy or clinical recommendations. The immunoassay comparison is more limited in precision (nÖ=Ö40 immunoassay positives), and confidence intervals for immunoassay-specific estimates are correspondingly wider and should be interpreted with caution.

## Results

### Participants

The final analytic sample comprised 119 male participants aged 18-40 years (mean (SD) = 28.8 (7.0) years). Of these, 84 (70.6%) resided in urban and 35 (29.4%) in rural kebeles. For the purposes of this research, key sample characteristics relevant to diagnostic accuracy interpretation are reported below. Both the immunoassay and HPLC were performed on the same urine sample collected at T2, and the ASR interview was conducted on the same day as urine collection, with T2 occurring 1-7 days following T1 baseline assessment. Immunoassay was conducted on raw urine at local health centres on the day of collection, whereas HPLC analysis was conducted at the Jimma University Laboratory of Drug Quality following refrigerated transport (mean (SD) = 4.9 (1.7) hours; range 1.6-8.2) and solid-phase extraction.

### Test Result Distributions

HPLC identified detectable urinary NE in 77 participants (64.7%) and no detectable NE in 42 (35.3%). The immunoassay returned a positive result (>300Öng/mL) for 40 participants (33.6%) and a negative result for 79 (66.4%). Binary ASR identified 91 khat users (76.5%) and 28 non-users (23.5%). Concerning the different use periods: 81 (89.0%) reported use within the past 0-2 days, and 10 (11.0%) reported their most recent use as 3 or more days prior to the interview.

No adverse events were reported in connection with the urine sampling, immunoassay procedure or the TLFB interview. Urine samples were collected using standard labelled containers; this is a non-invasive procedure, and no adverse events were associated with it. A small number of participants were initially unable to provide a urine sample at the time of the T2 appointment; these individuals were given time and opportunity to produce a sample during the appointment and were not excluded on this basis.

#### Baseline Demographics of Validation Subsample

#### Diagnostic Accuracy: Immunoassay vs. HPLC

Table 2 presents the 2×2 contingency table. The immunoassay produced zero false positives and 37 false negatives relative to HPLC.

**Table 1.**
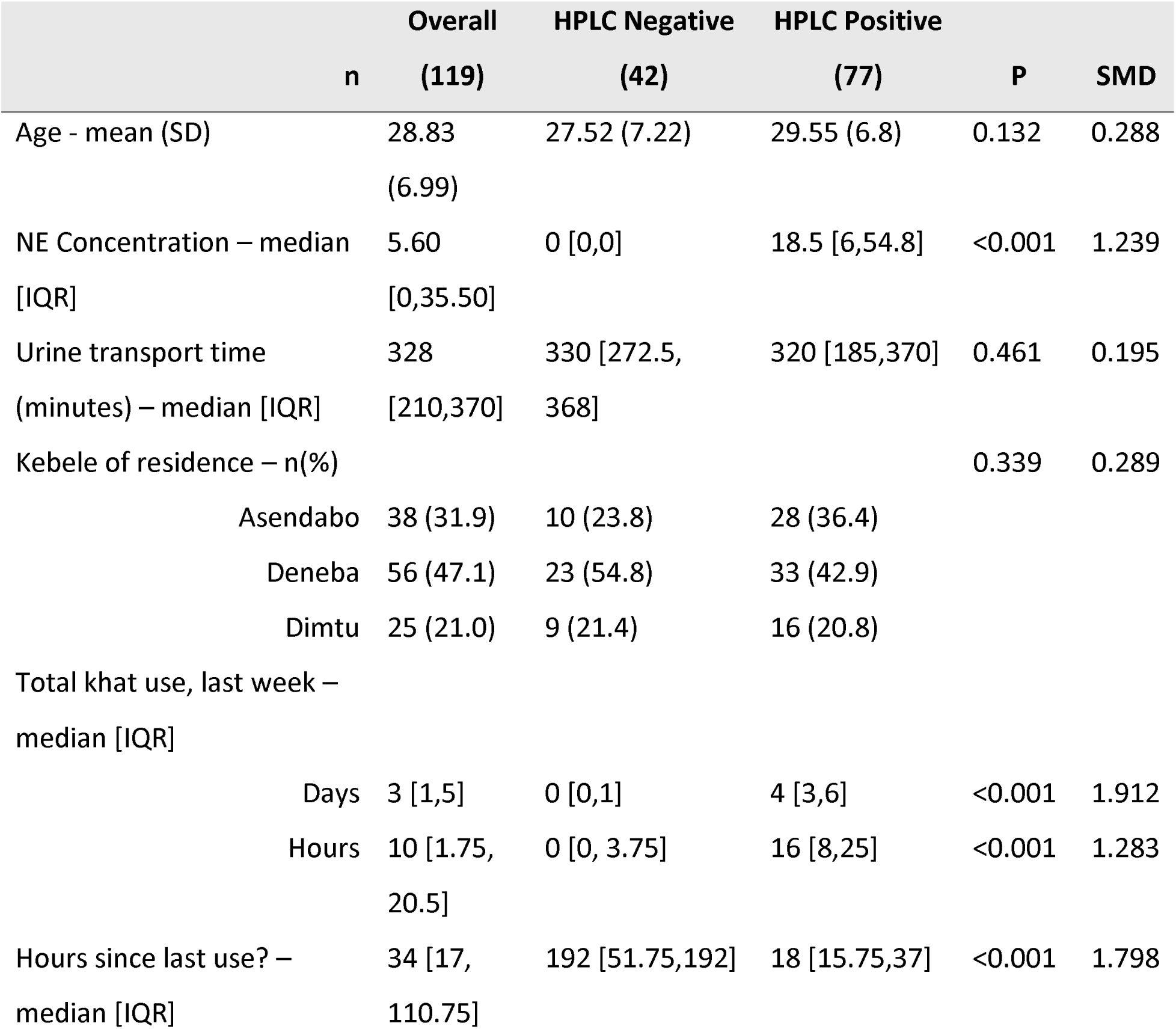
SMD = standardised mean difference. Continuous variables presented as mean (SD) or median [IQR]. Categorical variables presented as n (%).

|  | Overall<br>n<br>(119) | HPLC Negative<br>(42) | HPLC Positive<br>(77) | P | SMD |
| --- | --- | --- | --- | --- | --- |
| Age - mean (SD) | 28.83<br>(6.99) | 27.52 (7.22) | 29.55 (6.8) | 0.132 | 0.288 |
| NE Concentration – median<br>[IQR] | 5.60<br>[0,35.50] | 0 [0,0] | 18.5 [6,54.8] | <0.001 | 1.239 |
| Urine transport time<br>(minutes) – median [IQR] | 328<br>[210,370] | 330 [272.5,<br>368] | 320 [185,370] | 0.461 | 0.195 |
| Kebele of residence – n(%) |  |  |  | 0.339 | 0.289 |
| Asendabo | 38 (31.9) | 10 (23.8) | 28 (36.4) |  |  |
| Deneba | 56 (47.1) | 23 (54.8) | 33 (42.9) |  |  |
| Dimtu | 25 (21.0) | 9 (21.4) | 16 (20.8) |  |  |
| Total khat use, last week –<br>median [IQR] |  |  |  |  |  |
| Days | 3 [1,5] | 0 [0,1] | 4 [3,6] | <0.001 | 1.912 |
| Hours | 10 [1.75,<br>20.5] | 0 [0, 3.75] | 16 [8,25] | <0.001 | 1.283 |
| Hours since last use? –<br>median [IQR] | 34 [17,<br>110.75] | 192 [51.75,192] | 18 [15.75,37] | <0.001 | 1.798 |

**Table 2.** 2×2 contingency table: amphetamine immunoassay (300Öng/mL) vs. HPLC (nÖ=Ö119). TPÖ= true positive; FNÖ= false negative; FPÖ= false positive; TNÖ= true negative.

|  | Immunoassay<br>Positive | Immunoassay<br>Negative | Total |
| --- | --- | --- | --- |
| HPLC Positive | 40 (TP) | 37(FN) | 77 |
| HPLC Negative | 0 (FP) | 42 (TN) | 42 |
| Total | 40 | 79 | 119 |

**Table 3.** Diagnostic accuracy metrics: immunoassay vs. HPLC. Benchmark: ≥0.80 sensitivity and specificity (pre-specified).

| Diagnostic Metric | Estimate [95% CI] |
| --- | --- |
| Sensitivity | 0.519 [0.403, 0.635] |
| Specificity | 1.0 [0.916, 1.0] |
| PPV | 1.0 [0.912, 1.0] |
| NPV | 0.532 [0.416, 0.645] |
| Overall Accuracy | 0.689 [0.598, 0.771] |
| Cohen's Kappa | 0.433 (weak) |

The immunoassay achieved perfect specificity (1.0) and PPV (1.0), confirming that all immunoassay-positive results corresponded to true khat users. However, sensitivity was substantially below the 0.80 benchmark (0.52), indicating that approximately half of biologically confirmed khat users were missed. The NPV of 0.53 means that a negative immunoassay result had little diagnostic value, only marginally better than chance at ruling out use. Kappa indicated weak agreement (0.43).

Among the 77 HPLC-positive participants, mean urinary NE concentration was significantly higher in true positives (immunoassay-positive; mean (SD) = 60.7 (44.8) µg/mL, n = 40) than in false negatives (immunoassay-negative; mean (SD) = 11.7 (18.3) µg/mL, n = 37; Welch’s t[52.5] = −6.36, p < 0.001, Cohen’s d = 1.41). This is consistent with immunoassay false negatives being concentrated among participants with lower NE concentrations.

#### Diagnostic Accuracy: Binary ASR vs. HPLC

Table 4 presents the 2×2 contingency table for binary ASR versus HPLC. ASR produced 17 false positives and 3 false negatives.

**Table 4.** 2×2 contingency table: binary ASR vs. HPLC (nÖ=Ö119).

|  | ASR Positive | ASR Negative | Total |
| --- | --- | --- | --- |
| HPLC Positive | 74 (TP) | 3 (FN) | 77 |
| HPLC Negative | 17 (FP) | 25 (TN) | 42 |
| Total | 91 | 28 | 119 |

**Table 5.**
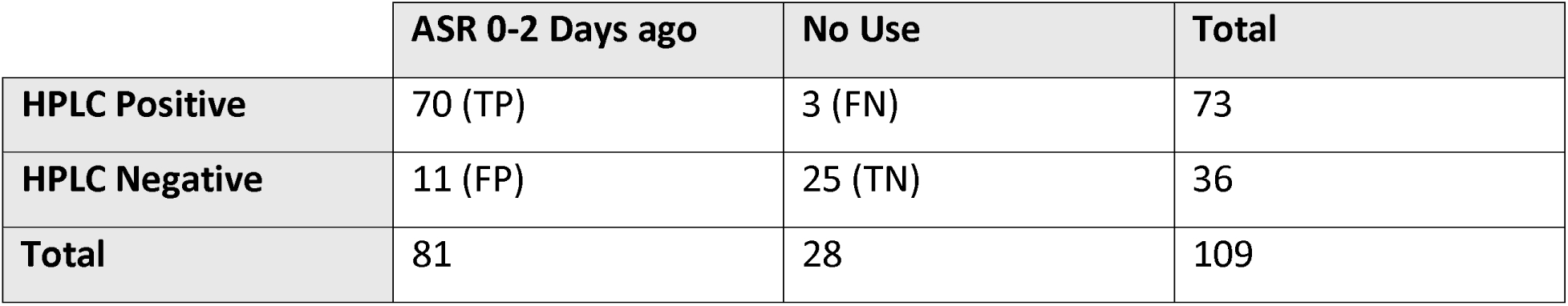
Contingency table: ASR 0-2 days vs. HPLC (nÖ=Ö109; participants with use only on days 3-7 excluded). Time-stratified 3+ days ASR vs. HPLC (pre-specified secondary analyses)

**Table 6.** Contingency table: ASR 3+ days vs. HPLC (nÖ= 38 ; participants with use only on days 0-2 excluded). ASR diagnostic accuracy by recall window

|  | ASR 3+ Days ago | No Use | Total |
| --- | --- | --- | --- |
| HPLC Positive | 4 (TP) | 3 (FN) | 7 |
| HPLC Negative | 6 (FP) | 25 (TN) | 36 |
| Total | 10 | 28 | 38 |

Binary ASR met the 0.80 sensitivity benchmark (0.96) against HPLC but not the specificity benchmark (0.60). NPV was high (0.89), supporting the reliability of negative ASR classifications (see Table 7).

**Table 7.** Time-stratified ASR vs. HPLC: comparison across recall windows. Note: wide 95% CIs for the 3-7 day window (nÖ=Ö38) limit interpretation.

| Metric | Binary ASR (n=119) | ASR 0-2 Days (n=109) | ASR 3-7 Days (n=38) |
| --- | --- | --- | --- |
| Sensitivity | 0.961 [0.890, 0.992] | 0.959 [0.885, 0.991] | 0.571 [0.184, 0.901] |
| Specificity | 0.595 [0.433, 0.744] | 0.694 [0.519, 0.837] | 0.806 [0.625, 0.925] |
| PPV | 0.813 [0.718, 0.887] | 0.864 [0.770, 0.930] | 0.400 [0.122, 0.738] |
| NPV | 0.893 [0.718, 0.977] | 0.893 [0.718, 0.977] | 0.893 [0.718, 0.977] |
| Overall Accuracy | 0.832 [0.752, 0.894] | 0.872 [0.794, 0.928] | 0.763 [0.598, 0.886] |
| Kappa | 0.602 (moderate) | 0.692 (moderate) | 0.324 (minimal) |

Among the 77 HPLC-positive participants, the three false negatives (ASR-negative despite detectable NE) had a mean (SD) NE concentration of 8.7 (4.7) µg/mL compared with 38.3 (42.8) µg/mL in the 74 true positives (Welch’s t[29.7] = −5.23, p < 0.001). However, given the extremely small false-negative group (n = 3), this result should be interpreted as descriptive rather than inferential.

Time-stratified 0-2 day ASR vs. HPLC (pre-specified secondary analyses)

Restricting to use within 0-2 days improved specificity from 0.60 to 0.69, PPV from 0.81 to 0.86, and kappa reached its strongest agreement (0.69), while preserving sensitivity (0.96 to 0.96). The 3-7 days window showed markedly reduced sensitivity (0.57) with wide confidence intervals, consistent with most urinary NE having cleared beyond the five-day detection limit.

In the 0-2 day window, the same pattern held: false negatives (n = 3, mean NE = 8.7 (4.7) µg/mL) had lower concentrations than true positives (n = 70, mean NE = 40.0 (43.4) µg/mL; Welch’s t[31.9] = −5.35, p < 0.001), though the small cell size limits interpretation. In the 3-7 day window, NE concentrations were comparably low in both true positives (n = 4, mean = 9.0 (7.3) µg/mL) and false negatives (n = 3, mean = 8.7 (4.7) µg/mL; t[4.9] = −0.08, p = 0.94), consistent with substantial metabolite clearance beyond the detection window regardless of classification status.

### Concordance: ASR vs. Immunoassay

Table 8 presents the 2×2 contingency table for ASR versus immunoassay as a field reference.

**Table 8.**
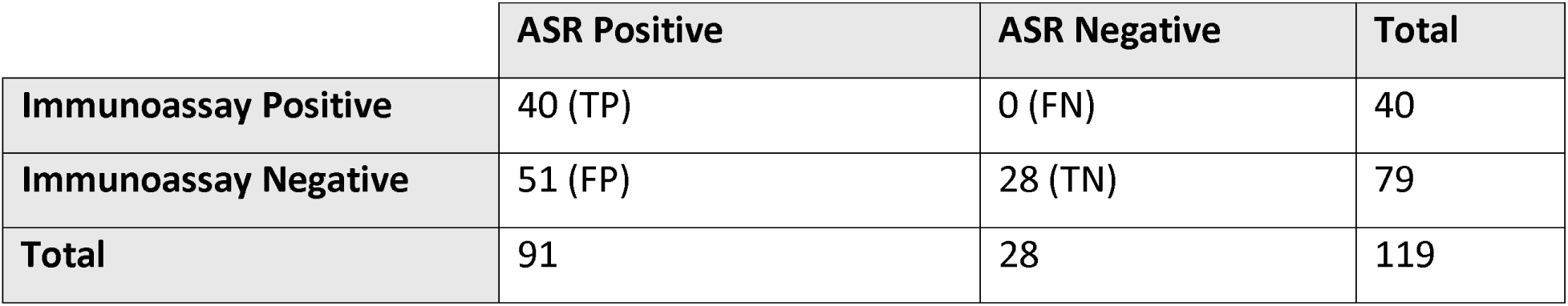
2×2 contingency table: binary ASR vs. immunoassay (nÖ=Ö119).

**Table 9.** Diagnostic accuracy metrics: binary ASR vs. immunoassay.

| Concordance Metric | Estimate [95% CI] |
| --- | --- |
| Positive Percent Agreement (PPA) | 1.0 [0.912, 1.0] |
| Negative Percent Agreement (NPA) | 0.354 [0.250, 0.470] |
| Overall Percent Agreement (OPA) | 0.571 [0.477, 0.662] |
| Cohen's Kappa | 0.270 (minimal) |

ASR achieved perfect PPA (1.0) against the immunoassay: every immunoassay-positive participant had also self-reported khat use, and no participant who denied use tested positive on the immunoassay. However, NPA was low (0.35), with 51 of 91 ASR-positive participants (56%) testing negative on the immunoassay. Critically, the HPLC data make it likely that this disagreement reflects immunoassay false negatives (not ASR false positives): of these 51 discordant participants, 34 (66.7%) had detectable urinary NE by HPLC (mean (SD) = 12.0 (19.1) µg/mL), consistent with genuine recent khat use, while the remaining 17 had no detectable NE, suggesting use reported outside the biological detection window.

Time-stratified ASR vs. immunoassay analyses showed modest improvements when restricted to 0-2 days (NPA 0.40 vs.Ö0.35; kappa 0.32 vs.Ö0.27, both minimal), with the 3-7 day window producing near-zero kappa (0.14, none).

Among the 91 ASR-positive participants, immunoassay-concordant participants (n=40) reported substantially more hours of khat use in the preceding 48 hours than immunoassay-discordant participants (n= 51): mean (SD) 8.06 (6.14) vs. 4.71 (4.19) hours (Welch’s t = −2.95, df = 65.9, p=0.004, Cohen’s d = 0.65). The same pattern held for total hours across the full recall week: mean (SD) 24.08 (19.49) vs. 14.4 (12.15) hours (Welch’s t = −2.75, df = 61.9, p=0.008, Cohen’s d = 0.61). Concordant participants reported approximately 70% more use in the preceding 48 hours, consistent with a dose-dependent relationship between consumption intensity and urinary NE accumulation above the 300 ng/mL immunoassay threshold.

## Discussion

This study systematically evaluated three field-feasible khat-use measures against laboratory HPLC in a prospective validation sub-study nested within a large Ethiopian male cohort. The key findings are: (1) the immunoassay’s 300Öng/mL cutoff produces low sensitivity (0.52) for detecting any khat use, despite perfect specificity; (2) binary TLFB ASR shows high sensitivity (0.96) and NPV (0.89) against HPLC; and (3) restricting ASR to the 0-2 day recall window improves specificity and elevates kappa to its strongest agreement with the reference standard (0.69), suggesting that pharmacokinetic misalignment, not recall bias, is the primary source of ASR-HPLC discordance.

The immunoassay’s specificity confirms its utility as a rule-in test: a positive result reliably identifies khat use, though it specifically reflects high-dose khat use or very recent use rather than any use. However, sensitivity of 0.52 means it fails to detect one in every two biologically confirmed khat users. Prior pharmacokinetic studies suggest that cathinone from khat chewing produces peak urinary NE concentrations that vary substantially by dose, chewing duration, and individual metabolic rate. The immunoassay’s low sensitivity reflects its detection chemistry and cutoff rather than poor assay performance. Because the test is calibrated to amphetamine and registers khat-derived cathinone metabolites only through antibody cross-reactivity, the 300 ng/mL threshold sits well above the NE concentrations many khat users produce, so true users fall below the detection cutoff. The nal von minden 300 ng/mL strip was in fact the best-performing of the assay evaluated in systematic pretesting; the limitation is therefore one of cutoff calibration for this analyte, not of this product relative to its peers, and a lower khat-calibrated cutoff would likely improve sensitivity substantially. These findings should not be generalized to amphetamine immunoassays as a class or to khat-calibrated alternatives.

The quantitative NE analysis provides direct mechanistic evidence for the immunoassay’s poor sensitivity. Among HPLC-positive participants, those correctly identified by the immunoassay (true positives) had mean NE concentrations more than five times higher than those missed (false negatives): 60.7 versus 11.7 µg/mL (Cohen’s d = 1.41). This large effect size is consistent with the 300 ng/mL cutoff systematically excluding participants whose khat use produced genuine but lower-concentration urinary NE whether due to lower dose, shorter chewing duration, or more distant timing of last use. The finding strengthens the case for validating a lower immunoassay cutoff calibrated to the pharmacokinetic profile of khat-derived NE.

The amphetamine immunoassay used here detects cathinone and its metabolites only through cross-reactivity, not direct binding, meaning higher urinary analyte concentrations are required to register positive than for equivalent amphetamine exposure. Since these data were originally collected, two complementary developments address this limitation. Lower-cutoff amphetamine immunoassays are now commercially available and would partially compensate for the affinity gap without changing the underlying detection mechanism. More directly, cathine-calibrated immunoassays (designed for d-nor pseudoephedrine) have entered the market, including NarcoCheck NCE-CAT-1S with a 150 ng/mL cathine-specific cutoff and manufacturer-reported 99% reliability^23^. By directly targeting a khat-relevant metabolite, these assays should detect the sub-threshold concentrations characteristic of the false-negatives observed here at comparable per-test cost. Together, these developments suggest the sensitivity limitations documented in this study are addressable in future validation work without sacrificing field deployment feasibility.

The ASR comparisons reinforce this interpretation from the opposite direction. The three false negatives in the binary ASR-HPLC comparison had uniformly low NE concentrations (mean = 8.7 µg/mL), suggesting these participants may have been infrequent or light users for whom recall was less salient, though the small cell size (n = 3) precludes firm conclusions. In the 3-7 day time-stratified window, NE concentrations were uniformly low (∼9 µg/mL) in both true positives and false negatives, consistent with near-complete metabolite clearance. This convergence of low NE values across classification groups in the extended recall window provides further evidence that pharmacokinetic timing, rather than reporting bias, drives the ASR-HPLC discordance observed in the binary comparison.

The intended role of ASR in this context is as a replacement measure for biological testing in large-scale community surveillance. By this criterion, binary ASR performs acceptably: sensitivity and overall accuracy both exceed 0.80, and the high NPV (0.89) makes it particularly useful for correctly identifying non-users, a critical function in epidemiological studies estimating exposure-outcome associations. The moderate specificity (0.60) is a known limitation of past-week binary ASR, attributable primarily to pharmacokinetic rather than behavioural misalignment as demonstrated by the time-stratified analyses. For studies where biological validation will be conducted, using the 0-2 day ASR window as the primary exposure variable is recommended to optimise concordance with biological detection.

Characterisation of discordant pairs across all three of the diagnostic comparisons displays a coherent pattern. The 17 false positives are most plausibly explained by reported use occurring outside the NE urinary detection window (i.e., use 6-7 days prior when metabolites may have cleared) rather than deliberate overreporting, given that the TLFB asks about specific days rather than a global yes/no. The three false negatives (denied use, detectable NE) suggest a small degree of underreporting, possibly reflecting social desirability or forgetting in infrequent users. The immunoassay’s 300Öng/mL threshold similarly accounts for the poor ASR-immunoassay agreement, not overreporting by participants. Taken together, the discordant pair analyses across all three comparisons converge on a single conclusion: misclassification in this sample is driven primarily by analytical sensitivity limitations of the immunoassay and pharmacokinetic timing of metabolite clearance, not by systematic reporting bias in the ASR. This conclusion is further supported by the ASR-immunoassay comparison: no participant denied khat use while testing positive on the immunoassay, suggesting that underreporting is negligible in this population. This may reflect the normative acceptability of khat use in this rural, predominantly Oromo setting, where male chewing is a routine social practice and carries little stigma. Underreporting could be more pronounced in settings where khat use is less socially sanctioned so the negligible underreporting observed here should not be assumed to generalize to populations where the behaviour is stigmatized.

### Implications for practice

This diagnostic accuracy study serves as the foundational measurement validation component of a larger investigation examining the relationship between khat use and psychiatric outcomes in the GGFRC cohort. Accurate classification of the khat use exposure variable is a prerequisite for any subsequent analysis of khat-related health outcomes: misclassification of users as non-users or vice versa will attenuate observed associations and undermine causal inference. The findings reported here therefore directly inform which measurement approach should be used to classify exposure status in that parent investigation. Based on these findings, the following recommendations are made:

1. The standard 300Öng/mL amphetamine immunoassay (nal von minden, Drug-Screen AMP test) is not suitable as a primary any-use classification tool in this population. Its sensitivity of 0.52 means it systematically misclassifies approximately half of true khat users as non-users, which would produce substantial non-differential misclassification bias in exposure-outcome analyses. However, its perfect specificity and PPV make it well-suited as a confirmatory marker of heavy or recent high-dose use which is a potentially important distinction given evidence of dose-dependent relationship between khat consumption intensity and health outcomes. Researchers should therefore consider deploying it not as a general use detector, but as a targeted measure of substantial exposure when biological confirmation of heavy use is the specific research or clinical objective. These limitations are specific to this test at this cutoff and should not be generalized to amphetamine immunoassay as a class or khat-calibrated alternatives.
2. TLFB binary ASR (past-week use) is an acceptable primary exposure measure for large-scale khat epidemiology, with sensitivity (0.96) and overall accuracy (0.83) both exceeding the 0.80 benchmark against HPLC. Its high NPV (0.89) makes it particularly reliable for correctly identifying non-users. Specificity limitations (0.60) should be acknowledged and interpreted in light of the pharmacokinetic evidence that most false positives reflect metabolite clearance rather than overreporting.
3. The relative importance of sensitivity versus specificity and NPV versus PPV, depends on the intended use of the measure. For epidemiological studies estimating exposure-outcome associations, high sensitivity and NPV are prioritised to mitigate the threat of misclassifying users as non-users, which would attenuate observed effects. For clinical screening or confirmatory purposes, where ruling in exposure with confidence matters more, the immunoassay’s perfect PPV makes it the more appropriate tool despite its low sensitivity. Researchers should also select measures based on the consequences of misclassification in their specific context.
4. Where biological validation is available alongside ASR, data collection should be timed to capture ASR within 0-2 days of urine collection to maximise concordance with the biological detection window.
5. In field settings where formal TLFB administration is not feasible, a single binary question “have you used khat in the past 48 hours?” would capture the majority of biologically confirmed recent users.
6. The immunoassay can serve as an add-on confirmatory measure alongside ASR. A positive immunoassay result can be used with confidence to confirm khat-user status, and more specifically high khat use, but a negative immunoassay result should not be used to reclassify an ASR-positive participant as a non-user given the immunoassay’s low sensitivity at the 300Öng/mL cutoff.

## Limitations

Generalisability: The analytic sample comprised males aged 18-40 from a single region of Ethiopia. Findings may not generalise to women, older adults, populations in other khat-using regions, or settings with different khat varieties (which may produce different alkaloid concentrations).

Sample size and precision: The validation sample (nÖ=Ö119) was determined practically. While adequate for the primary comparisons, precision is limited for the 3-7 day stratified analysis (nÖ=Ö38, with only n = 7 HPLC-positive), where confidence intervals are wide. The immunoassay also produced only 40 positive results, limiting the precision of immunoassay-specific estimates.

Disentangling bias from pharmacokinetic misalignment: The study design cannot fully separate social desirability bias from pharmacokinetic window misalignment as explanations for ASR-HPLC discordance. However, the convergent evidence from the time-stratified analyses strongly implicates pharmacokinetic rather than behavioural mechanisms as the primary driver (i.e. diagnostic metrics improved markedly in the 0-2 day window).

Single validation timepoint: The validation was conducted at T2 only. Measurement performance may vary across seasons, which requires further investigation.

Sampling limitation: At baseline (T1) of the 1,100 originally recruited participants 25.9% (n=259) declined consent. If these participants systematically differed from those who consented in their khat use patterns, the prevalence estimates may have been negatively affected, however diagnostic metrics are less susceptible to this form of selection bias.

Dose and use-pattern assessment: The measures evaluated here classify khat use as a binary or time-stratified exposure and do not capture dose, within-session intensity, or phytochemical properties of the consumed khat. The TLFB does record hours of use per day, used here as a proxy for consumption intensity, but quantitative dose was not incorporated into the diagnostic accuracy framework, which is a limitation given evidence of dose-dependent relationship between consumption intensity and biological detectability.

## Other Information

Ethical approval and consent: This study received ethical approval from the Jimma University Institutional Review Board (Ref: RPGC/546/2014 and Ref: IHRPGD/2069/2017) and the Ludwig Maximilian University of Munich Institutional Review Board. Written informed consent was obtained from all participants in their preferred language (Afaan Oromo or Amharic).

Community engagement involved briefings with kebele leaders, employment of local enumerators from the community, and a commitment to disseminate results through community meetings. Participants reporting severe mental health symptoms were referred to local health services with their consent.

Study protocol: For further information regarding study design, data collection procedures, please contact the last author: Michael Odenwald, University of Konstanz, Konstanz, Germany. For inquiries regarding the analysis plan for this validation study, please contact the corresponding author: Hugh Atkinson, Dalhousie University, Halifax, Nova Scotia, Canada.

## Data availability

De-identified individual participant data and the analytic code supporting the findings of this study are available from the corresponding author upon reasonable request and subject to data sharing agreement with the parent study investigators at the University of Konstanz and Jimma University. Public deposition is not currently feasible due to ongoing analyses within the parent cohort and ethical restrictions on the original consent.

## Author contributions

Conceptualization: HA, KA, MO. Methodology: HA, KA, MM, MA, MO, SSu, ST, SSt. Investigation: MO, KA, MW, VM, ZM, SSu, MS. Formal analysis: HA. Data curation: HA, KA, MO, LO. Visualization: HA. Resources: KA, MO. Validation: KA, MM, MA, SSt, SSu, ZM, ST, MO, LO. Supervision: KA, MO, MM, MA, SSt, SSu, ZM. Project administration: KA, MO. Funding acquisition: KA, MO, TGS, MM. Writing – original draft: HA. Writing – review & editing: KA, MO, MM, MA, SSt, LO. All authors reviewed and approved the final version of the manuscript.

## Supporting information

STARD Checklist

## Data Availability

De-identified individual participant data and the analytic code supporting the findings of this study are available from the corresponding author upon reasonable request and subject to a data-sharing agreement with the parent-study investigators at the University of Konstanz and Jimma University. Public deposition is not currently feasible due to ongoing analyses within the parent cohort and ethical restrictions on the original consent.

## Acknowledgements

We thank the sixteen local enumerators of the GGFRC, the laboratory staff at the Jimma University Laboratory of Drug Quality, the GGFRC field team, and the study participants, without whom this study would not have been possible to conduct. Furthermore, we thank Heike Riedke for training local health center staff in urine handling and testing. We thank Dr. Fasil Tessema for his contribution and overall support for the project.

## Appendices

Appendix A: TLFB Grid

**Figure 3.**
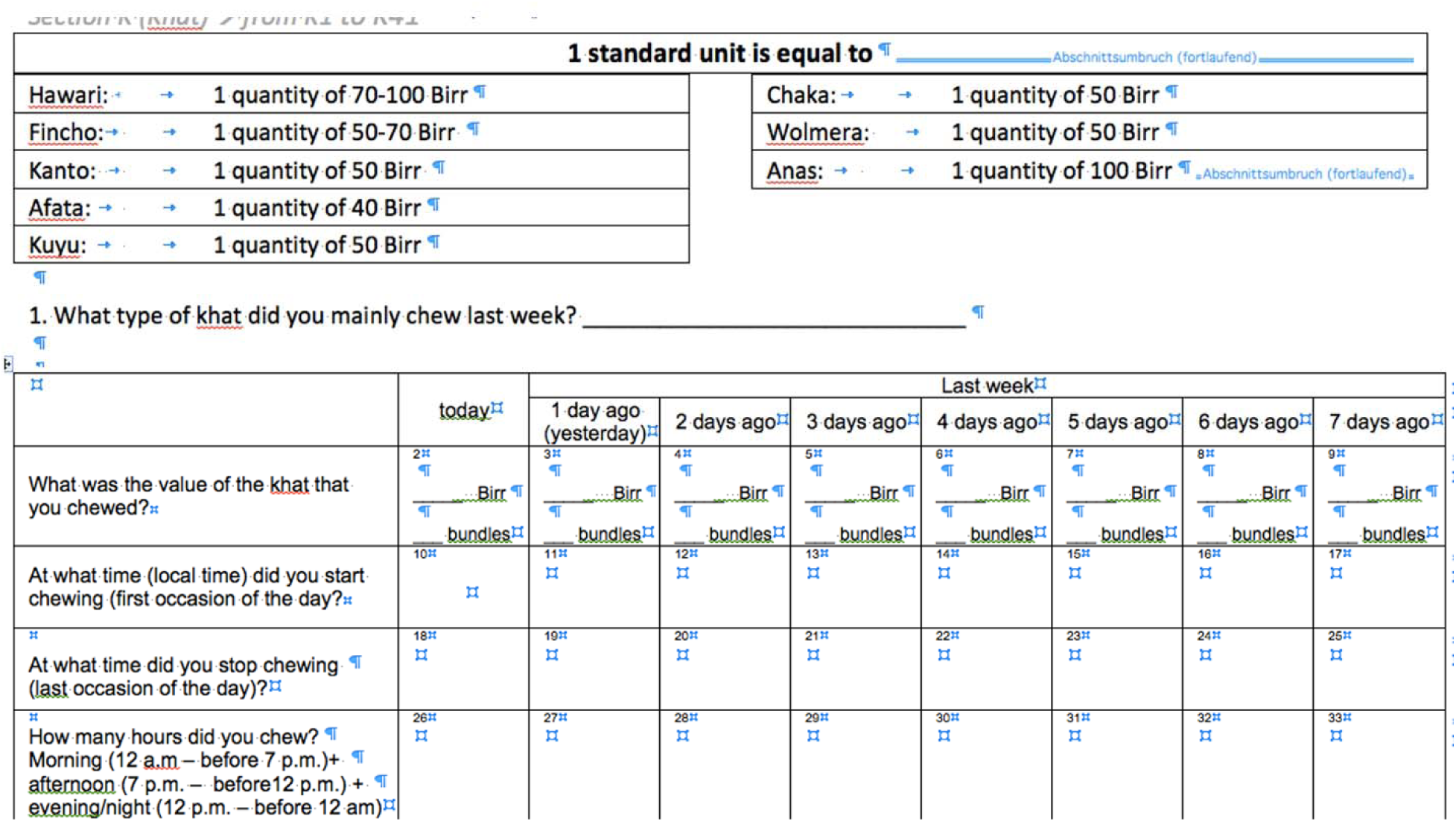
**TLFB Grid. Note: Taken from the original questionnaire used during data collection.**

## References

1. Ahmed A, Ruiz MJ, Kadosh KC, Patton R, Resurrección DM. Khat and neurobehavioral functions: A systematic review. PLoS ONE. 2021. doi:10.1371/journal.pone.0252900

2. Silva B, Soares J, Rocha-Pereira C, MLaděnka P, Remião F, Researchers OBOTO. Khat, a Cultural Chewing Drug: A Toxicokinetic and Toxicodynamic Summary. Toxins. 2022. doi:10.3390/toxins14020071

3. Alemu WG, Zeleke TA, Takele WW, Mekonnen SS. Prevalence and risk factors for khat use among youth students in Ethiopia: Systematic review and meta-analysis, 2018. Annals of General Psychiatry. 2020. doi:10.1186/s12991-020-00265-8

4. Olani AB, Gerbaba M, Getnet M, Soboka M, Decorte T. Is chewing khat associated with mental health disorders? A scoping review of the content and quality of the current evidence base. Subst Abuse Treat Prev Policy. 2023 Jun 27;18(1):39. doi:10.1186/s13011-023-00545-y

5. Toennes SW, Harder S, Schramm M, Niess C, Kauert GF. Pharmacokinetics of cathinone, cathine and norephedrine after the chewing of khat leaves. British Journal of Clinical Pharmacology. 2003;56(1). doi:10.1046/j.1365-2125.2003.01834.x

6. Saitman A, Park HD, Fitzgerald RL. False-positive interferences of common urine drug screen immunoassays: A review. Journal of Analytical Toxicology. 2014. doi:10.1093/jat/bku075

7. Toennes SW, Kauert GF. Excretion and Detection of Cathinone, Cathine, and Phenylpropanolamine in Urine after Kath Chewing. Clinical Chemistry. 2002 Oct 1;48(10):1715–9. doi:10.1093/clinchem/48.10.1715

8. Bedada W, De Andrés F, Engidawork E, Hussein J, LLerena A, Aklillu E. Effects of Khat (Catha edulis) use on catalytic activities of major drug-metabolizing cytochrome P450 enzymes and implication of pharmacogenetic variations. Sci Rep. 2018 Aug 24;8(1):12726. doi:10.1038/s41598-018-31191-1

9. Adorjan K, Odenwald M, Widmann M, Tesfaye M, Tessema F, Toennes S, et al. Khat use and occurrence of psychotic symptoms in the general male population in Southwestern Ethiopia: evidence for sensitization by traumatic experiences. World Psychiatry. 2017 Oct;16(3):323. doi:10.1002/wps.20470 PubMed PMID: 28941092; PubMed Central PMCID: PMC5608818.

10. Cohen JF, Korevaar DA, Altman DG, Bruns DE, Gatsonis CA, Hooft L, et al. STARD 2015 guidelines for reporting diagnostic accuracy studies: explanation and elaboration. BMJ Open. 2016 Nov 14;6(11):e012799. doi:10.1136/bmjopen-2016-012799 PubMed PMID: 28137831; PubMed Central PMCID: PMC5128957.

11. Dehning S, Odenwald M, Girma E, Tessema F, Tesfaye M, Mekonnen Z, et al. The Impact of Lifestyle on Mental Health among Young Men in the Gilgel Gibe Field Research Center, Ethiopia. Ludwig Maximilian University; 2013.

12. Univeristy J. Gilgel Gibe Field Research Center [Internet]. Available from: https://ju.edu.et/gilgel-gibe-field-research-center/

13. Network I. Gilgel HDSS [Internet]. Available from: https://www.indepth-network.org/Profiles/Gilgel%20HDSS.pdf

14. Wood EA, Case SJ, Collins SL, Stark H, Wilfong T. From traditional to transactional: exploration of khat use in Ethiopia through an interpretative phenomenological analysis. BMC Public Health. 2024 Jul 15;24(1):1887. doi:10.1186/s12889-024-19357-1

15. Toennes SW. Khat – analysis of biofluids and pharmacokinetics [Conference Presentation]. Conference Presentation presented at: The First European Khat Research Program Conference. 2013 Sep 26; Frankfurt am Main, Germany.

16. nal von minden GmbH. nal von minden Drug-Screen® AMP urine test strip: product information and instructions for use [Internet]. Version 1.04. 2025 Jan 30;(Ref. 101016). Available from: https://nal-vonminden.com/deu/drug-screen-amp-urin-teststreifen.htmL

17. Sobell LC, Sobell MB. Timeline Follow-Back. In: Measuring Alcohol Consumption. 1992. doi:10.1007/978-1-4612-0357-5_3

18. Chen R, Cogburn M. Drug Testing. StatPearls [Internet]. 2025 Dec 1. Available from: https://www.ncbi.nlm.nih.gov/books/NBK459334/

19. Moeller KE, Lee KC, Kissack JC. Urine Drug Screening: Practical Guide for Clinicians. Mayo Clinic Proceedings. 2008 Jan;83(1):66–76. doi:10.4065/83.1.66

20. McHugh ML. Interrater reliability: The kappa statistic. Biochemia Medica. 2012;22(3). doi:10.11613/bm.2012.031

21. Center for Devices and Radiological Health. Statistical Guidance on Reporting Results from Studies Evaluating Diagnostic Tests: Guidance for Industry and FDA Staff [Internet]. Rockville, MD: U.S. Food and Drug Administration; 2007. Available from: https://www.fda.gov/regulatory-information/search-fda-guidance-documents/statistical-guidance-reporting-results-studies-evaluating-diagnostic-tests-guidance-industry-and-fda

22. Santos GM, Strathdee SA, El-Bassel N, Patel P, Subramanian D, Horyniak D, et al. Psychometric properties of measures of substance use: a systematic review and meta-analysis of reliability, validity and diagnostic test accuracy. BMC Med Res Methodol. 2020 May 7;20(1):106. doi:10.1186/s12874-020-00963-7 PubMed PMID: 32380951; PubMed Central PMCID: PMC7203822.

23. PharmaDrugTest.com [Internet]. [cited 2026 Apr 29]. Urine test for Cathine (for Khat/Kat/Qat/Chat). Available from: https://www.pharmadrugtest.com/urine-drug-tests/96-cathine-urine-test-khat-chat.htmL

